# Associations of Physical Activity and Heart Rate Variability from a Two-Week ECG Monitor with Cognitive Function and Dementia: the ARIC Neurocognitive Study

**DOI:** 10.1101/2024.03.01.24303633

**Authors:** Francesca R. Marino, Hau-Tieng Wu, Lacey Etzkorn, Mary R. Rooney, Elsayed Z. Soliman, Jennifer A. Deal, Ciprian Crainiceanu, Adam P. Spira, Amal A. Wanigatunga, Jennifer A. Schrack, Lin Yee Chen

## Abstract

**BACKGROUND:** Low physical activity (PA) measured from accelerometers and low heart rate variability (HRV) measured from short-term ECG recordings are associated with worse cognitive function. Wearable long-term ECG monitors are now widely used. These monitors can provide long-term HRV data and, if embedded with an accelerometer, they can also provide PA data. Whether PA or HRV measured from long-term ECG monitors is associated with cognitive function among older adults is unknown.

**METHODS:** Free-living PA and HRV were measured simultaneously over 14-days using the Zio^®^ XT Patch among 1590 participants in the Atherosclerosis Risk in Communities Study [aged 72-94 years, 58% female, 32% Black]. Total amount of PA was estimated by total mean amplitude deviation (TMAD) from the 14-day accelerometry raw data. HRV indices (SDNN and rMSSD) were measured from the 14-day ECG raw data. Cognitive factor scores for global cognition, executive function, language, and memory were derived using latent variable methods. Dementia or mild cognitive impairment (MCI) status was adjudicated. Linear or multinomial regression models examined whether higher PA or higher HRV was cross-sectionally associated with higher factor scores or lower odds of MCI/dementia. Models were adjusted for demographic and medical comorbidities.

**RESULTS:** Each 1-unit higher in total amount of PA was significantly associated with 0.30 higher global cognition factor scores (95% CI: 0.16-0.44), 0.38 higher executive function factor scores (95% CI: 0.22-0.53), and 62% lower odds of MCI (OR: 0.38, 95% CI: 0.22-0.67) or 75% lower odds of dementia (OR: 0.25, 95% CI: 0.08-0.74) versus unimpaired cognition. Neither HRV measure was significantly associated with cognitive function or dementia.

**CONCLUSIONS:** PA derived from a 2-week ECG monitor with an embedded accelerometer was significantly associated with higher cognitive test performance and lower odds of MCI/dementia among older adults. By contrast, HRV indices measured over 2 weeks were not significantly associated with cognitive outcomes. More research is needed to define the role of wearable ECG monitors as a tool for digital phenotyping of dementia.

**CLINICAL PERSPECTIVE:** *What Is New?:*

- This cross-sectional study evaluated associations between physical activity (PA) and heart rate variability (HRV) measured over 14 days from a wearable ECG monitor with cognitive function.
- Higher total amount of PA was associated with higher global cognition and executive function, as well as lower odds of mild cognitive impairment or dementia.
- HRV indices measured over 2 weeks were not significantly associated with cognitive outcomes.

*What Are the Clinical Implications?:*

- These findings replicate positive associations between PA and cognitive function using accelerometer data from a wearable ECG monitor with an embedded accelerometer.
- These findings raise the possibility of using wearable ECG monitors (with embedded accelerometers) as a promising tool for digital phenotyping of dementia.

## INTRODUCTION

Physical activity (PA) is a known modifiable risk factor for cognitive impairment and dementia.^1^ There is a growing interest in objectively measuring PA using accelerometers. These devices are small, non-invasive, and continuously capture free-living movement over multiple days.^2, 3^ This method has important advantages for measuring PA among older adult populations who typically spend most time sedentary or performing lighter intensity PA.^2^ There are many different brands of accelerometers currently used in research.^2, 3^ Yet, there are barriers to integrating these devices into clinical environments.^4^ Recent work from our group found that accelerometry data measured by the Zio^®^ XT Patch, a commonly used ambulatory electrocardiogram (ECG) monitor capable of continuous (i.e., uninterrupted) wear for up to 14 days, was highly correlated with ActiGraph wrist accelerometry data.^5^ To determine the potential utility of wearable ECG monitors with embedded accelerometers, it is important to evaluate associations between accelerometer-measured PA and important clinical outcomes, such as cognitive impairment. However, whether PA measured from these ECG monitors is related to cognitive function has not yet been established.

Heart rate variability (HRV) is defined as variation in successive heartbeats and lower HRV is associated with poor health outcomes.^6–8^ Previous studies among adults without cognitive impairment have shown lower HRV is associated with poorer global cognition,^9, 10^ memory,^9, 11^ language,^9^ attention,^9^ executive function,^9, 10^ visuospatial performance,^9^ and processing speed.^9^ Lower HRV is also related to lower cognitive test performance among those with Alzheimer’s disease^12^ and cognitive impairment.^8, 10^ This prior work is based on HRV measures from ECG recordings taken over several minutes in clinical settings or 24-hour recordings taken in the free-living environment. Longer ECG recordings might be more accurate in representing an individual’s HRV, particularly in response to the environment.^6, 13^ To date, no studies have evaluated associations between HRV from long-term ECG recordings and cognitive function.

Thus, we aimed to address these gaps by leveraging data from Zio^®^ XT Patch ECG and accelerometry data measured over 14 days in the free-living environment. First, we evaluated the cross-sectional association of total PA with cognitive function among older adults. Second, we examined the cross-sectional association between long-term HRV and cognitive function. We hypothesized that lower total PA and lower HRV are associated with lower cognitive test performance and higher odds of dementia than higher PA or HRV, respectively.

## METHODS

### Study Population

The Atherosclerosis Risk in Communities (ARIC) study is a prospective cohort study of 15792 community-dwelling mostly Black and White adults aged 45-64 years at enrollment. Participants were enrolled from Forsyth County, North Carolina; Jackson, Mississippi; Minneapolis, Minnesota; or Washington County, Maryland between 1987 and 1989.^14^ This analysis included participants without permanent atrial fibrillation who attended Visit 6 in 2016-2017 and wore the Zio^®^ XT Patch (iRhythm Technologies, San Francisco, California). Participants missing cognitive tests, dementia status, or covariate data were excluded (**Figure 1**). All study protocols were approved by the institutional review board at each study site, and all participants provided informed consent.

**Figure 1.**
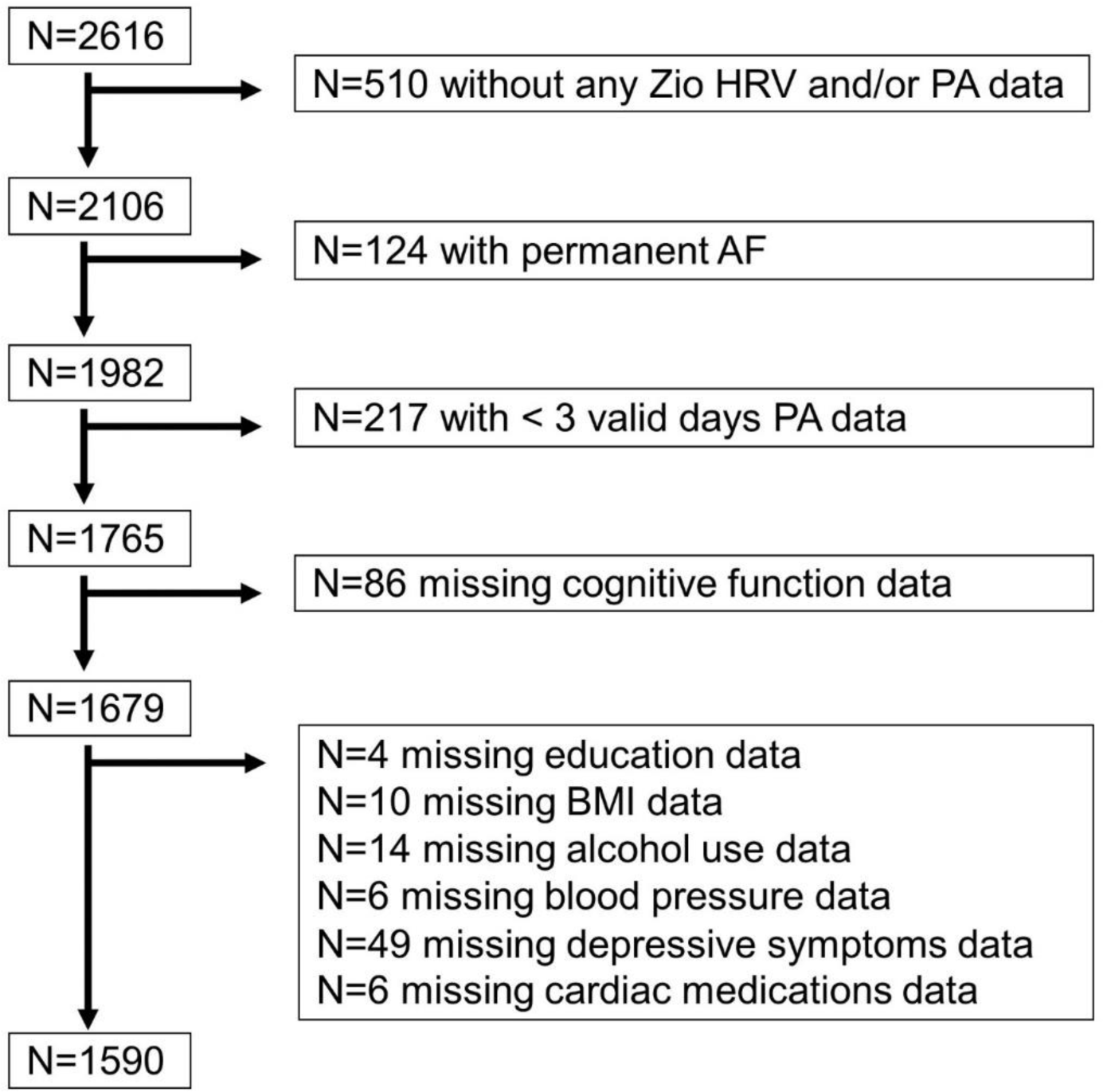
Flowchart of Sample Selection. Note: PA=physical activity; AF=atrial fibrillation; BMI=body mass index.

### Heart Rate Variability

HRV measures were derived from continuous ECG raw data over 14 days from the Zio^®^ XT Patch. The patch was placed on the participant’s chest and worn continuously until the end of data collection. After data collection, the participant mailed the patch to iRhythm Technologies Inc, for data processing. The primary HRV measures of interest included two HRV indices in the time domain: 1) the standard deviation of normal-to-normal RR intervals (SDNN); and 2) the root mean squared successive difference in normal-to-normal RR intervals (rMSSD). Lower SDNN and rMSSD are associated with poor health outcomes. ^6–8^ RR intervals are the time between successive normal heartbeats, SDNN represents the standard deviation between heartbeats, and rMSSD describes beat-to-beat variance.^6, 15^ SDNN and rMSSD were log transformed due to non-normal distributions and then analyzed continuously. Additional details of HRV data processing can be found in the supplemental materials (**Heart Rate Variability Data Processing**).

### Physical Activity

First, accelerometry raw data from the Zio^®^ XT Patch tri-axial accelerometer were summarized into minute-level mean amplitude deviation (MAD), a measure of movement intensity with gravitational units.^16^ MAD is strongly correlated with activity counts,^17^ heart rate,^16^ and PA intensity.^16^ A recent study also validated MAD from the Zio^®^ XT Patch against total activity counts measured using the ActiGraph wrist accelerometer.^5^ Second, minute-level MAD measurements were screened for periods of wear and non-wear based on the simultaneously measured ECG data. Third, days with more than 144 (10%) non-wear minutes were excluded, and participants with fewer than three valid days were excluded from further analysis. Finally, minute-level data were summarized into participant-level total mean amplitude deviation (TMAD), which is the daily total MAD averaged over all valid days. Higher TMAD can therefore be interpreted as higher levels of PA.^16, 17^ TMAD was natural log transformed due to non-normal distributions and then analyzed continuously (LTMAD). Processing of minute-level activity data was performed using the arctools R package.^18^

### Cognitive Test Factor Scores

Cognitive test performance was defined by factor scores for global cognition, executive function, memory, and language at Visit 6. Participants completed a neuropsychological test battery with tests administered by trained psychometrists.^19^ Executive function tests included Trail Making Test parts A and B^20^ and Digit Symbol Substitution Test.^21^ Memory tests included delayed word recall and logical memory from the Wechsler Memory Scale – Revised^22^ and incidental learning from the Wechsler Adult Intelligence Scale III.^23^ Language tests included semantic and phonemic fluency^24^ and Boston Naming tests.^25^ From these individual cognitive tests, cognitive factor scores for global cognition, executive function, memory, and language were derived using confirmatory factor analysis.^19^ Confirmatory factor analysis is a latent variable approach that uses all available cognitive test data to generate factor scores with a common scale while accounting for the common covariation among cognitive tests.^19^ Missing cognitive test data were addressed by using maximum likelihood with robust standard errors.^19^ Advantages of confirmatory factor analysis include: 1) use of all available test data; 2) allowing individual tests to have unequal weights; and 3) accounting for measurement error of individual tests.^19^ The full details of the cognitive factor score derivation in ARIC are described elsewhere.^19^

### Mild Cognitive Impairment or Dementia Status

Participants were classified as having dementia, mild cognitive impairment (MCI), or unimpaired cognition at Visit 6. Classifications were algorithmically based on criteria from the National Institute on Aging – Alzheimer’s’ Association workgroups^26, 27^ and 5^th^ Edition Diagnostic and Statistical Manual of Mental Disorders.^28^ This algorithm used information from the Mini-Mental State Exam,^29^ Clinical Dementia Rating Scale-Sum of Boxes,^30^ cognitive tests, and Functional Activities Questionnaire^31^ to define participants as having highly likely or probable unimpaired cognition, MCI, or dementia or having an indeterminate status.^32^

Participants were defined as having MCI by: at least one cognitive test < 1.5 standard deviations below the mean; cognitive test decline below the 10^th^ or 20^th^ percentiles for one or two tests, respectively; Clinical Dementia Rating Scale-Sum of Boxes score > 0.5 but ≤ 3; and Functional Activities Questionnaire score ≤ 5.^32^ Participants were defined as having dementia by: at least two cognitive tests < 1.5 standard deviations below the mean; cognitive test decline below the 10^th^ or 20^th^ percentiles for one or two tests, respectively; Clinical Dementia Rating Scale-Sum of Boxes score > 3; and Functional Activities Questionnaire score > 5.^32^ Mini-Mental State Examination scores < 21 for White participants or < 19 for Black participants also defined a dementia classification.^32^ Participants were classified as cognitively unimpaired (i.e., normal cognition) if they did not meet criteria for MCI or dementia.^32^ Data were reviewed by a panel of clinicians for participants classified as having MCI or dementia that was highly likely or probable or who had an uncertain status. The full details of MCI and dementia classifications in ARIC are described elsewhere.^32^

### Other Covariates

The following covariates were measured at Visit 1: age (years); sex (male, female); race/center; and education (grade school, high school without degree, high school graduate, vocational school, at least some college, or graduate/professional school). Covariates measured at Visit 6 included: smoking status (current, former, never); alcohol status (current, former, never); body mass index (BMI; kilograms/meters^2^); systolic and diastolic blood pressure (mmHg); history of diabetes; history of heart failure; depressive symptoms (Center for Epidemiologic Studies-Depression Scale score, which ranges from 0-60 with higher scores indicating higher depressive symptoms);^33^ and cardiac medications (beta-blocker, digoxin, calcium channel blocker, or anti-arrhythmic). Each medication was individually included as a covariate.

### Statistical Methods

To provide basic summaries of the analytic sample, participants were categorized into PA and HRV tertiles. Variables were summarized as tertile-specific means and standard deviations or counts and proportions, and differences between tertiles were tested using F-tests or chi-squared tests for continuous or categorical variables, respectively.

Separate linear regression models estimated the mean cross-sectional difference in global cognition, memory, executive function, or language factor scores per 1-unit increment in LTMAD, log SDNN, or log rMSSD. Separate multinomial logistic regression models estimated the odds of MCI or dementia versus unimpaired cognition per 1-unit increment in LTMAD, log SDNN, or log rMSSD. Diurnal curves of MAD over 24 hours were plotted by dementia status. The final adjusted models included age, sex, race/center, education, smoking, alcohol, systolic and diastolic blood pressure, BMI, diabetes, heart failure, depressive symptoms, and each cardiac medication.

The models described above were fit with additional sample exclusions or adjustments in sensitivity analyses. First, LTMAD, log SDNN, and log rMSSD were included in the same adjusted models to determine whether there was an independent effect of PA above and beyond HRV, or vice versa. Second, individuals with a history of stroke and/or non-permanent atrial fibrillation (i.e., intermittent atrial fibrillation) or who were taking beta-blockers or calcium channel blockers were excluded. Third, those with MCI or dementia were excluded from the cognitive test score models. All analyses were conducted using Stata statistical software version 17 (StataCorp, College Station, TX), and p-values < 0.05 were considered statistically significant.

## RESULTS

### Sample Characteristics

Of the n=2616 participants who attended ARIC Visit 6 and wore the Zio^®^ XT Patch, 1590 participants were included in the analysis (**Figure 1**). The overall sample characteristics of these 1590 participants are described in **Tables 1-2**. Participants were on average aged 78.8 (range: 72-94) years, 58% were female, and 32% were Black. Participants with higher PA were younger; less likely to be Black; and more likely to be male, current drinkers, and former smokers compared to those with lower PA. This group also had higher education; lower body mass index and systolic blood pressure; and lower prevalence of diabetes, heart failure, and depressive symptoms (**Table 1**). Participants with higher HRV were less likely to be Black, more likely to be male, had lower BMI, and lower prevalence of diabetes than those with lower HRV (**Table 2**).

**Table 1.**
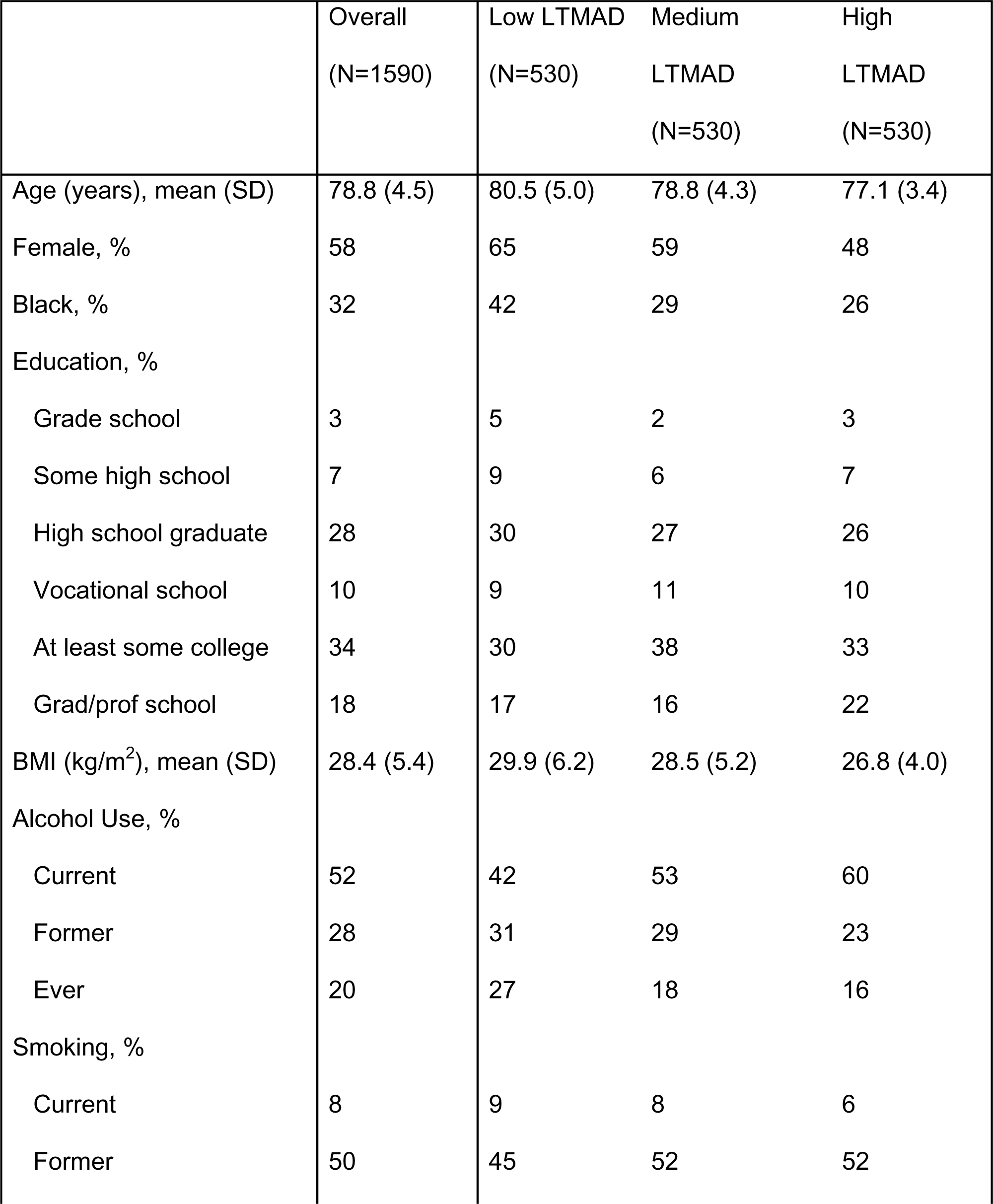
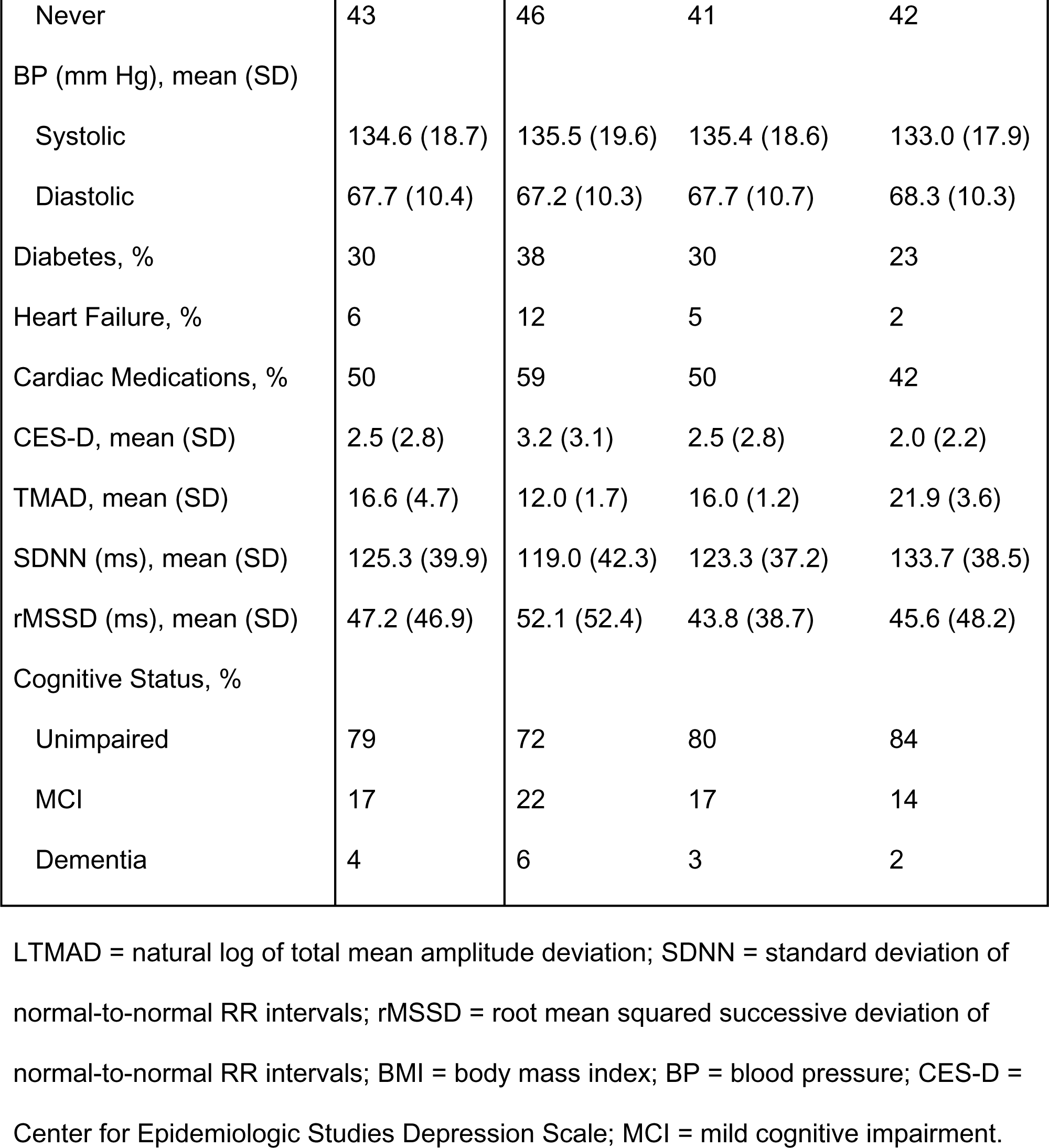
Sample Characteristics Overall and by Tertiles (Low, Medium, High) of LTMAD.

**Table 2.**
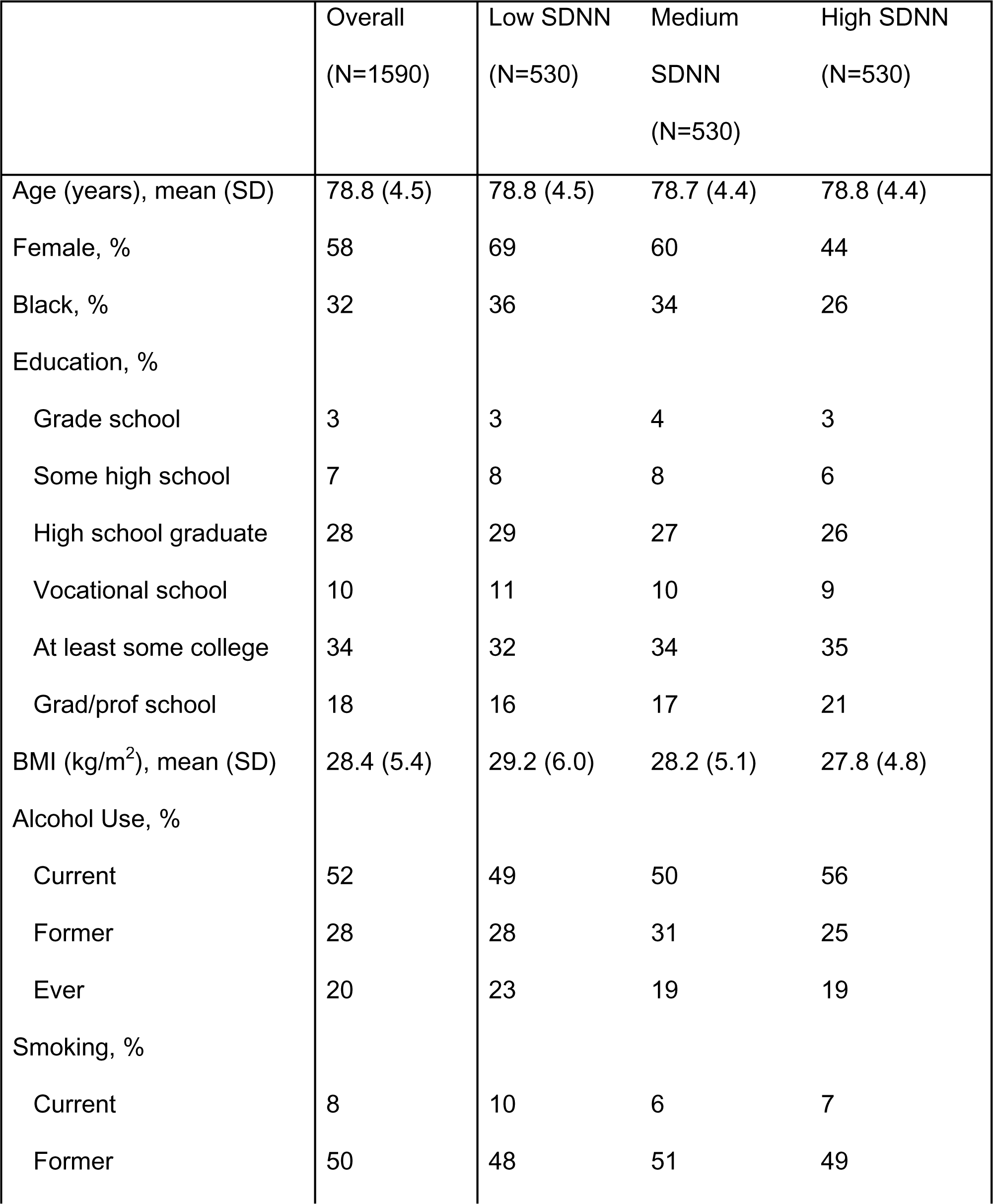
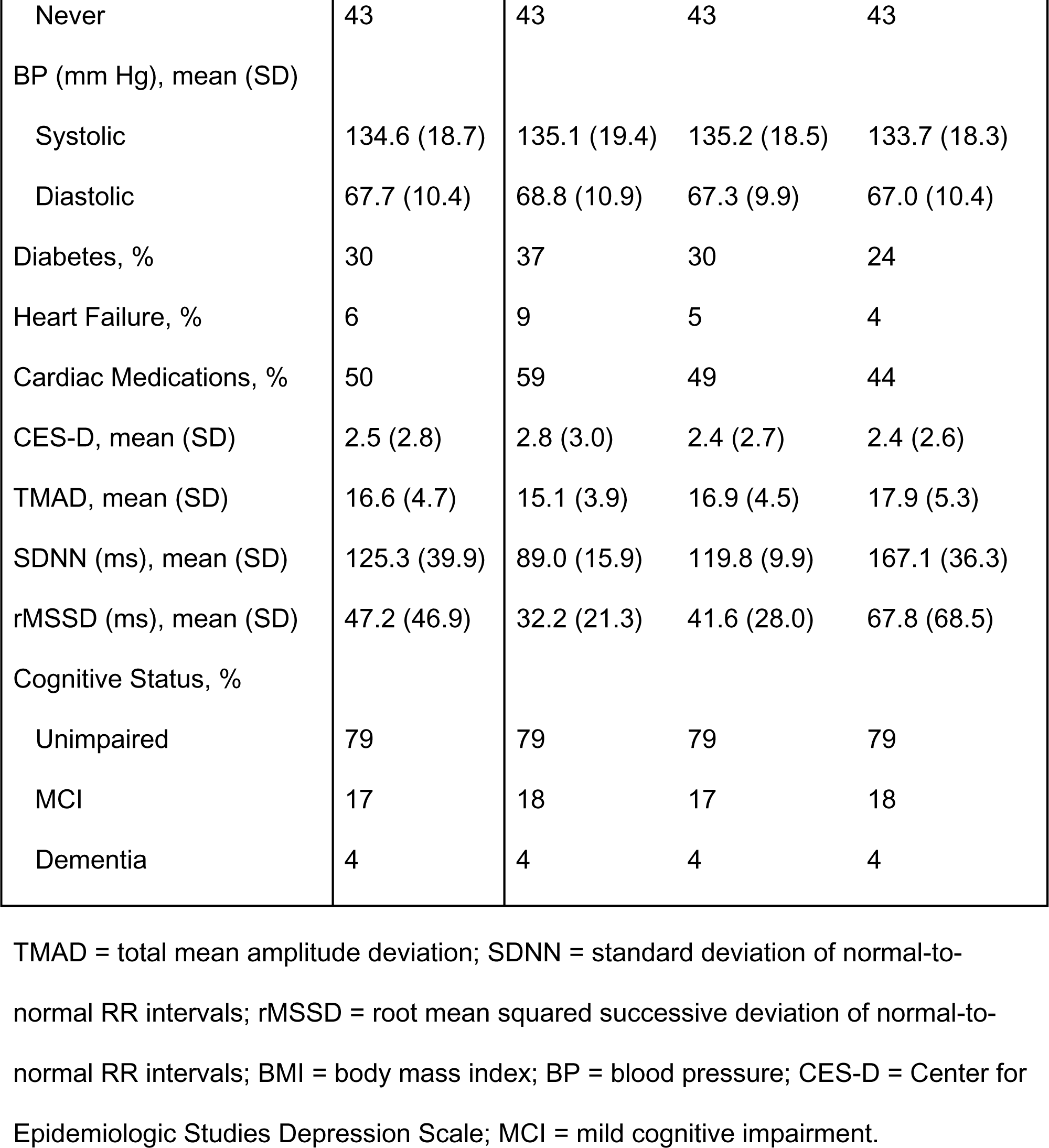
Sample Characteristics Overall and by Tertiles (Low, Medium, High) of log SDNN.

TMAD was weakly correlated with SDNN and with rMSSD (Pearson’s correlation = 0.22 or −0.08, respectively). The two HRV measures (i.e., SDNN and rMSSD) were moderately correlated (Pearson’s correlation = 0.48) (**Figure S1**).

### Physical Activity and Cognitive Function

In fully adjusted models, each 1-unit increment in LTMAD was significantly associated with 0.30 higher global cognition (95% CI: 0.16, 0.44) and 0.38 higher executive function factor scores (95% CI: 0.22, 0.53). There was a non-significant association with language score (0.15 higher language factor score per 1-unit increment in LTMAD [95% CI: −0.02, 0.32; p=0.079]) and no associations with memory factor scores (**Table 3**).

**Table 3.** Cross-Sectional Differences in Cognitive Test Scores by LTMAD, log SDNN, or log rMSSD Over 24-Hours.

| | Global<br>Cognition<br>$\beta$ (95% CI) | Executive Function<br>$\beta$ (95% CI) | Memory<br>$\beta$ (95% CI) | Language<br>$\beta$ (95% CI) |
| --- | --- | --- | --- | --- |
| LTMAD | **0.30**<br>**(0.16, 0.44)** | **0.38**<br>**(0.22, 0.53)** | 0.14<br>(-0.04, 0.32) | 0.15<br>(-0.02, 0.32) |
| Log SDNN | 0.03<br>(-0.24, 0.30) | 0.19<br>(-0.10, 0.48) | -0.21<br>(-0.56, 0.13) | -0.15<br>(-0.47, 0.17) |
| Log rMSSD | -0.12<br>(-0.2, 0.010) | -0.08<br>(-0.2, 0.06) | -0.16<br>(-0.3, 0.004) | -0.11<br>(-0.26, 0.05) |

Linear regression model adjusted for: age, sex, race/center, education, smoking, drinking, systolic and diastolic blood pressure, body mass index, diabetes, heart failure, depressive symptoms, and cardiac medications; N=1590.

When looking at diurnal curves of PA in **Figure 2**, participants without cognitive impairment (blue line) had the highest peak in activity. This group also had higher daytime activity than those with MCI (green line) or dementia (gray line). Participants with MCI and dementia had blunted diurnal patterns of activity, with those with dementia having the greatest attenuation in activity (**Figure 2**). In fully adjusted models, each 1-unit increment in LTMAD was associated with 62% lower odds of MCI (OR=0.38, 95% CI: 0.22, 0.67) or 75% lower odds of dementia (OR=0.25, 95% CI: 0.08, 0.74) versus unimpaired cognition (**Table 4**).

**Figure 2.**
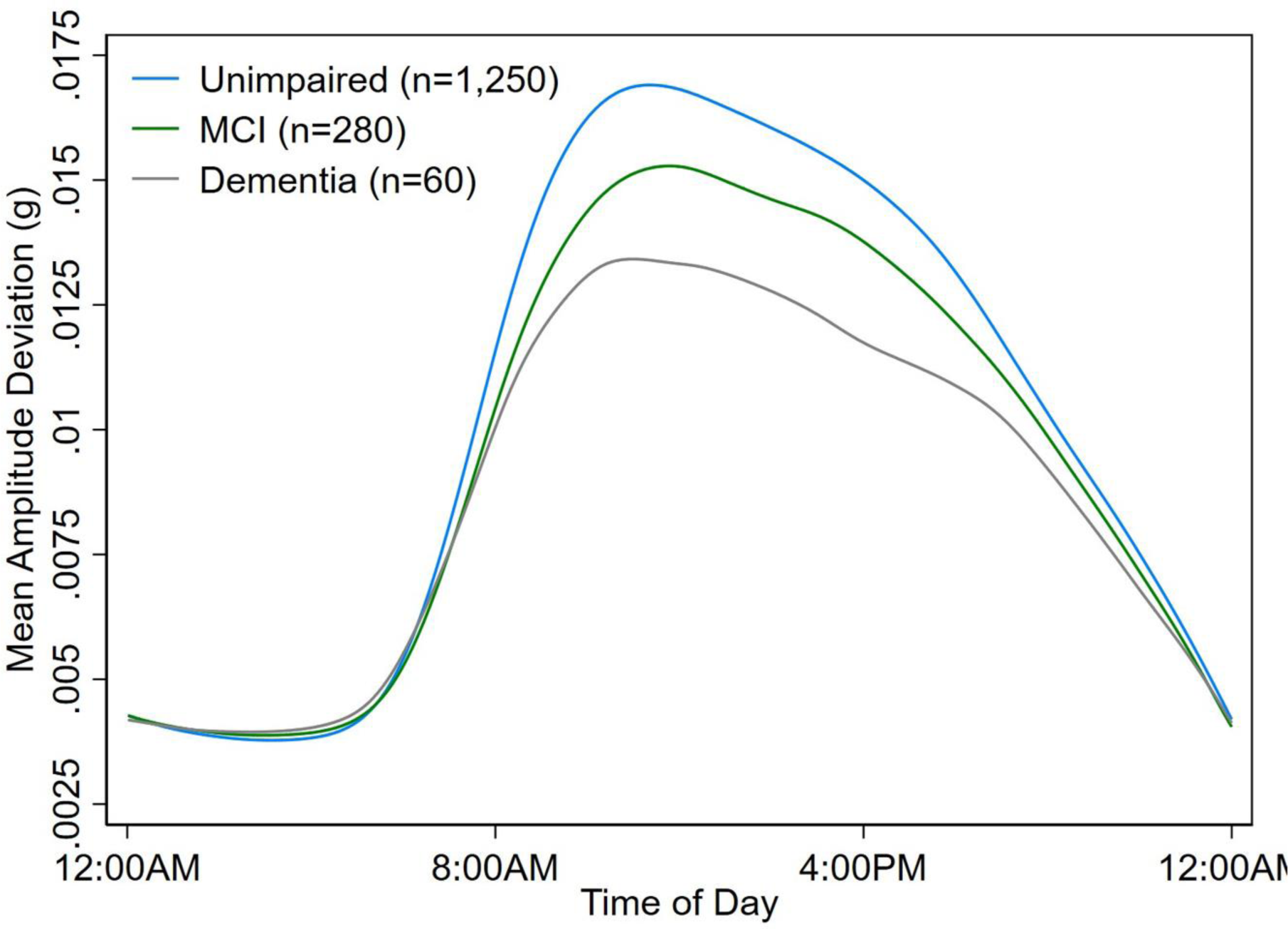
Diurnal Patterns of MAD by Dementia, Mild Cognitive Impairment, or Cognitively Unimpaired Status.

**Table 4.** Cross-Sectional Odds Ratio of MCI or Dementia Compared to Unimpaired Cognition by 1-Unit Higher in LTMAD, log SDNN, or log rMSSD Over 24-Hours.

|  | Unimpaired (N=1250) | Mild Cognitive Impairment (N=280)<br>OR (95% CI) | Dementia (N=60)<br>OR (95% CI) |
| --- | --- | --- | --- |
| LTMAD | REF | **0.38 (0.22, 0.67)<sup>a</sup>** | **0.25 (0.08, 0.74)** |
| Log SDNN | REF | 0.84 (0.30, 2.36) | 3.39 (0.40, 28.37) |
| Log rMSSD | REF | 1.40 (0.87, 2.27) | 1.24 (0.46, 3.35) |

Multinomial logistic regression model adjusted for: age, sex, race/center, education, smoking, drinking, systolic and diastolic blood pressure, body mass index, diabetes, heart failure, depressive symptoms, and cardiac medications; N=1590.
a. 1-unit higher in LTMAD (i.e., more physical activity) is associated with 62% lower odds of MCI relative to unimpaired cognition.

### Heart Rate Variability and Cognitive Function

There were no significant associations between log SDNN or log rMSSD and cognitive function. SDNN and rMSSD were not significantly associated with cognitive test performance (**Table 3**) nor with MCI or dementia (**Table 4**).

### Independent Effects of Physical Activity and Heart Rate Variability Measures

When including LTMAD, log SDNN, and log rMSSD together in the same model, associations for higher LTMAD with higher global cognition and executive function factor scores and lower odds of MCI or dementia remained significant. Further, 1-unit increment higher in LTMAD was associated with 0.15 higher memory factor scores (95% CI: 0.04, 0.33). Associations with log SDNN or log rMSSD and cognitive factor scores or dementia status remained non-significant (**Tables S1-2**).

### Exclusion of Stroke, Intermittent Atrial Fibrillation, Beta-Blockers, and Calcium Channel Blockers

When excluding N=170 participants with any history of stroke or intermittent atrial fibrillation and N=675 taking either beta-blockers or calcium channel blockers, associations for LTMAD with global cognition and executive function factor scores and dementia remained significant (**Table S3-4**). Further, each 1-unit increment in log SDNN was associated with 0.59 higher executive function factor scores (95% CI: 0.12, 1.07) (**Table S3**).

### Sensitivity Analyses Restricting by Cognitive Status

When excluding N=340 participants with MCI or dementia, associations between LTMAD and executive function factor scores remained robust, but global cognition findings were attenuated (p=0.09). Associations with log SDNN or log rMSSD and cognitive factor scores remained non-significant (**Table S5**).

## DISCUSSION

In a community-based sample of older adults, higher free-living PA derived from an accelerometer embedded in a wearable 2-week ECG monitor was cross-sectionally associated with higher cognitive test performance and lower odds of MCI/dementia. By contrast, time domain measures of HRV derived from 2-week ECG raw data were not associated with cognitive performance and MCI/dementia. These findings raise the possibility of using wearable ECG monitors (with embedded accelerometers) as a tool to predict dementia. These findings also challenge previously reported associations between lower HRV and poorer cognitive function based on short-term heart rate data.

Higher PA measured using Zio^®^ XT Patch accelerometry data was cross-sectionally associated with higher executive function and global cognition factor scores, as well as lower odds of MCI or dementia. These findings were largely robust across multiple sensitivity analyses. This is consistent with prior research documenting higher PA is associated with better cognitive performance ^34^ and lower risk of dementia,^1^ as well as research reporting that individuals living with MCI/dementia exhibit differences in objectively measured PA patterns.^35, 36^ Potential mechanisms explaining these associations include higher PA leading to increased brain neurotrophic factors^34, 37^ and cerebral blood flow^34, 37^ or decreased inflammation,^34^ stress overactivity,^34^ and cardiovascular risk factors.^38^ In addition, it is important to acknowledge that emerging evidence suggests the associations between cognition and PA might be bidirectional.^39^ However, we are unable to establish claims about temporality from this cross-sectional study. While this work suggests potential utility of long-term wearable ECG monitors with an embedded accelerometer for routinely measuring free-living PA, future longitudinal studies are needed to determine whether these data can be used to predict cognitive decline or dementia.

In our analyses, HRV was not related to cognitive test performance or odds of MCI/dementia. Previous work using short-term measurements of HRV suggests higher values of certain indices are associated with better cognition, but other studies report null results.^8, 9, 40, 41^ The non-significant findings in our study might be due to the measurement of free-living HRV over a longer ECG monitoring period of 14 days, which more accurately represent an individual’s HRV than shorter ECG recordings of several minutes to 24 hours.^13^ Further, our study focused on only two time-domain measures that are relatively constant over time.^6, 42^ Frequency-domain measures (i.e., high frequency, low frequency, etc.) which divide total HRV into certain frequency categories and represent different physiological systems^9^ might be more strongly related to cognitive function. Future studies are needed to evaluate whether other patterns of long-term HRV are related to cognitive function.

In sensitivity analyses excluding those with a history of stroke, atrial fibrillation, beta-blockers, or calcium channel blockers, we found that higher HRV, specifically SDNN, was associated with higher executive function. These results align with studies finding associations between higher HRV and better executive function.^9, 11, 40, 41, 43^ HRV is influenced by both components of the autonomic nervous system^6^ and represents an individual’s ability to adapt to changes in their environment (i.e., faster heart rate with stressor).^9^ Certain diseases and medications can impact HRV, including stroke,^44^ atrial fibrillation,^45^ and cardiac medications.^46, 47^ Having a history of stroke or atrial fibrillation is also associated with cognitive decline and risk of dementia.^48, 49^ As such, these factors might have hidden (i.e., confounded) the association between HRV and cognitive function when analyzing the whole sample. The mechanisms linking HRV with cognitive function should be evaluated in future work, while also considering potential differences in associations by certain cardiovascular diseases or medications.

This study suggests the possible utility of wearable ECG monitors with embedded accelerometers as a tool to assess cognitive function and predict dementia. This has important public health implications as many wearable ECG monitors are already commonly used in the clinical setting. Further, the measure of PA used in this study was a simple summary of total amount of activity, which adds to the feasibility of using these data. We had previously shown that this measure of total amount of PA was highly correlated with total activity counts from the ActiGraph accelerometer.^5^ Although the summary measures of HRV were not related to cognitive function in this study, it is possible that more detailed metrics of long-term HRV are associated with cognitive function. Specifically, it might be necessary to examine HRV by time-of-day. Higher daytime HRV is associated with positive health outcomes,^6–8^ whereas higher overnight HRV is linked with negative health outcomes.^50^ It might also be important to consider whether bouts of high HRV but low PA, or vice versa, are informative of cognitive function. This could include combining minute-level ECG and accelerometry data to generate time-varying profiles of PA and HRV. Beyond exploring additional HRV measures, replication of these findings in longitudinal cohorts with diverse samples is needed, as the findings from this cross-sectional study do not inform whether PA measured from this ECG monitor can predict cognitive decline or dementia.

### Limitations and Strengths

It is important to consider these results in the context of the study’s limitations. First, we are not able to establish temporality from these cross-sectional analyses. There is a possibility for reverse causation, where those with lower cognitive function might be less active and therefore have lower HRV. Second, we do not have information available on the context of activities participants were performing, which would help interpret the PA and HRV data. Third, there is possible selection bias, as participants must have had both Zio^®^ XT Patch and cognitive function data to be included in this analysis. If individuals with dementia or low cognitive test performance and worse PA or HRV did not wear the ECG device, then would expect this bias to be conservative (i.e., attenuated findings).

This study also has strengths. First, the ARIC study is a well-characterized cohort with a large sample of White and Black older adults. Second, ARIC conducts a comprehensive neuropsychological test battery and rigorous ascertainment of dementia, allowing us to examine associations across multiple cognitive domains and by dementia status. Third, we evaluated PA and HRV measured by a common clinical device in the free-living environment over 14-days.

### Conclusions

Higher free-living PA measured by a 2-week wearable continuous ECG monitor is positively associated with cognitive performance and MCI/dementia. By contrast, long-term time-domain measures of HRV are not associated with cognitive performance or MCI/dementia. Our findings suggest the potential of wearable continuous ECG monitors as a promising tool for digital phenotyping of dementia.

## Data Availability

Data are available from the ARIC Study Coordinating Center for investigators with an approved manuscript proposal form.

## ACKNOWLEDGEMENTS

The Atherosclerosis Risk in Communities Study is carried out as a collaborative study supported by National Heart, Lung, and Blood Institute contracts (75N92022D00001, 75N92022D00002, 75N92022D00003, 75N92022D00004, 75N92022D00005). The ARIC Neurocognitive Study is supported by U01HL096812, U01HL096814, U01HL096899, U01HL096902, and U01HL096917 from the NIH (NHLBI, NINDS, NIA and NIDCD). The authors thank the staff and participants of the ARIC study for their important contributions.

